# Validity of Dried Blood Spot Testing for Sexually Transmitted and Blood-Borne Infections: A Narrative Systematic Review

**DOI:** 10.1101/2023.04.06.23288121

**Authors:** François Cholette, Simone Périnet, Bronwyn Neufeld, Maggie Bryson, Jennifer Macri, Kathryn M. Sibley, John Kim, S. Michelle Driedger, Marissa L. Becker, Paul Sandstrom, Adrienne F. A. Meyers, Dana Paquette

## Abstract

**Objective:** Testing for human immunodeficiency virus (HIV) and hepatitis C virus (HCV) using dried blood spot (DBS) specimens has been an integral part of bio-behavioural surveillance in Canada for almost two decades. A systematic review was conducted to assess the current evidence regarding the validity of sexually transmitted and blood-borne infection (STBBI) testing using DBS specimens.

**Methods:** A literature search was conducted using a peer-reviewed search strategy. Eligibility criteria included studies reporting use of DBS specimens for STBBI testing in populations 15 years of age or older. The intervention of interest was either commercially available or “in-house” tests used to detect STBBI from DBS specimens. Studies that reported a measure of validity such as sensitivity, specificity, positive and negative predictive values were eligible for inclusion. Quality of studies and risk of bias were assessed using the QUADAS-2 tool.

**Results:** A total of 6,706 records were identified. Of these records, 169 full-text articles met the criteria for inclusion. The STBBI with the most articles reporting a measure of validity for testing on DBS was HIV (*n*=73), followed by HCV (*n*=63), HBV (*n*=33), syphilis (*n*=7), HAV (*n*=5), HSV (*n*=5), HTLV (*n*=3), and HPV (*n*=1). The majority of studies reported high sensitivity (≥90%) and specificity (≥90%). However, the quality of the studies varied greatly. No evidence was found on the validity of chlamydia and gonorrhoea testing using DBS specimens.

**Conclusion:** Our findings support the validity of DBS testing for STBBI surveillance where sufficient evidence was available, but validity is highly dependent on thorough method development and validation.

## Introduction

Dried blood spot (DBS) specimens consist of blood spotted and dried on filter paper. They have been used for biological sampling in clinical and research settings for nearly a century (Gruner, Stambouli, & Ross, 2015; Malsagova et al., 2020). Compared to conventional biological specimens like plasma or serum, DBS offers several advantages, including high acceptability among research participants (Landy et al., 2022) and compatibility with self-collection (Takano et al., 2018). DBS specimens are also stable at room temperature and do not require refrigeration during transportation, providing opportunities for biological sampling in remote and isolated communities (Lim, 2018). DBS sampling is typically deployed in resource-constrained settings for infectious disease diagnostics (Sherman, Stevens, Jones, Horsfield, & Stevens, 2005), surveillance (Buckton, 2008), and patient monitoring (Chevaliez & Pawlotsky, 2018), but it also has the potential to be an effective tool for infectious disease surveillance in high resource settings like Canada given the country’s numerous northern, remote, and isolated communities scattered across challenging geography. DBS sampling has also been shown to have a high acceptability as an approach of screening for sexually transmitted and blood-borne infections (STBBIs) in communities across Canada (Landy et al., 2022; Young et al., 2022).

DBS sampling was thrust into the spotlight during the COVID-19 pandemic, as it became routine for surveillance and epidemiological studies while limiting face-to-face interactions and overcoming healthcare staff shortages by allowing individuals to reliably collect their samples at home (Miesse, Collier, & Grant, 2022). For example, Statistics Canada’s Canadian COVID-19 Antibody and Health Survey (CCAHS) relied on DBS self-collection to assess COVID-19 seroprevalence nationally (www23.statcan.gc.ca/imdb/p2SV.pl?Function=getSurvey&SDDS=5339). As a result, lessons learned from implementing large-scale DBS sampling for SARS-CoV-2 surveillance could inform STBBI surveillance moving forward.

Diagnostic assays have shown excellent clinical performance on DBS specimens. Most validation studies have been performed for HIV screening, but more recent studies have demonstrated the feasibility of using DBS specimens for the diagnosis of viral hepatitis (Tuaillon et al., 2010). However, it is difficult to ascertain if DBS are a suitable specimen for STBBI surveillance considering the many experimental conditions (i.e. DBS punching and elution protocols), combinations of reference and index tests, and study populations. Special consideration is also needed for key populations where dual-infections are more prevalent, including HCV/HIV co-infection among people who inject drugs and HIV/syphilis co-infection in men who have sex with men (Gobran, Ancuta, & Shoukry, 2021; Roberts & Klausner, 2016). The presence of dual-infections is important to consider as it may have the potential to impact assay performance (McArdle, Turkova, & Cunnington, 2018). However, most validation studies focus on a single STBBI and therefore performance metrics are presented for a single index test, pathogen, and population, making it difficult to draw broader conclusions on the validity of using DBS specimens for STBBI surveillance.

The objective of this systematic review is to compile data on measures of validity to ascertain if the current literature supports the use of DBS, in the context of surveillance, for the detection of the following STBBI: HIV-1, HIV-2, hepatitis viruses (A, B, and C), herpes simplex virus (type 1 and 2), human T-cell lymphotropic virus (type 1 and 2), human papilloma virus, chlamydia (*Chlamydia trachomatis*), gonorrhoea (*Neisseria gonorrhoeae*), and syphilis (*Treponema pallidum*). Measures of validity include sensitivity, specificity, positive predictive values (PPV), negative predictive values (NPV), limit of quantification (LOQ), and/or limit of detection (LOD). Additionally, we aim to identify factors which may influence test performance.

## Methods

### Information Sources

Peer-reviewed original research was identified by searching Excerpta Medica dataBASE (EMBASE), Medical Literature Analysis and Retrieval System Online (Ovid MEDLINE) and Elsevier Scopus. Grey literature was identified by searching key websites including Public Health Ontario, BC Centre for Excellence in HIV/AIDS, Canadian AIDS Treatment Information Exchange (CATIE), Institut national d’excellence en santé et services sociaux, Open Grey, Public Health Agency of Canada, American Society of Microbiology, Infectious Disease Society of America, American Society of Virology, International AIDS Society, International Society for Sexually Transmitted Diseases Research, Conference on Retroviruses and Opportunistic Infections (CROI), International AIDS Conference, Centre for Disease Control (CDC), Public Health England, European Centre for Disease Prevention and Control, World Health Organization (WHO) and the French National Agency for AIDS Research (ANRS).

### Search Strategy

Literature searching was conducted under the guidance of a librarian (Janice Linton, University of Manitoba) using a peer-reviewed search strategy (McGowan et al., 2016). The search strategy consisted of both controlled vocabulary such as the National Library of Medicine’s MeSH (medical subject heading) and keywords (Supplementary File 1). Retrieval was limited to human populations, English and French language documents, and results were not limited by publication date up to May 2022.

### Eligibility Criteria

Populations were eligible if they provided DBS specimens tested for the following STBBIs: human immunodeficiency virus type 1 and 2 (HIV-1 and HIV-2), hepatitis virus A, B, and C (HAV, HBV, and HCV), human T-cell lymphotropic virus type 1 and 2 (HTLV-1 and HTLV-2), human papilloma virus (HPV), *Chlamydia trachomatis* (chlamydia), *Neisseria gonorrhea* (gonorrhoeae) and *Treponema pallidum* (syphilis). Studies were eligible if they were conducted on populations 15 years of age or older, regardless of socio-demographic characteristics and setting.

The intervention of interest was either commercially available or in-house tests used to detect STBBIs from DBS specimens. Inclusion criteria for commercial tests were limited to 3**^rd^** generation or greater enzyme immunoassays (EIAs) and nucleic acid tests, since older HIV testing methodologies (i.e., 1**^st^** and 2**^nd^** generation EIAs) are likely no longer manufactured (Alexander, 2016), and therefore are irrelevant.

The intervention was compared to either commercially available or in-house tests used to detect STBBIs from “gold-standard” biological specimens (ex: whole blood, plasma and serum) used for routine STBBI testing. Commercial tests were limited to 3**^rd^** generation or greater EIAs and nucleic acid tests.

Included studies had to report measures of the intervention’s validity such as sensitivity, specificity, positive predictive values (PPV), negative predictive values (NPV), limit of quantification (LOQ) and/or limit of detection (LOD).

### Exclusion Criteria

Peer-reviewed or grey literature were excluded if (1) the pathogen of interest was not included in the list of STBBIs mentioned above, (2) measures of the testing methodology’s validity were not reported, (3) biological specimens other than blood were collected on filter paper, (4) DBS were used to measure adherence to pre-exposure prophylaxis and/or anti-retroviral medication, (5) DBS were analyzed for the purpose of investigation of drug resistance or genotyping, (6) participants were under 15 years of age, (7) the intervention was out of scope (i.e., blood dried on matrix other than filter paper), (8) the work was not original research, or (9) the reports were in a language other than English or French.

### Document Screening

Titles and abstracts were initially screened by one reviewer (FC or BN) with 10% peer-reviewed by a second reviewer (SP or FC). A full-text review was then conducted of potential articles for inclusion by one reviewer (FC or BN) with 10% being peer-reviewed by a second reviewer (SP or FC). Disagreements were resolved by a third reviewer (MB).

### Data Extraction

Data was collected from each document on the inclusion list (e.g., study population, sampling method, sample size, index test, reference test, STBBI, DBS preparation method, sensitivity, specificity, accuracy, PPV, NPV, LOQ and/or LOD) and entered into a standardized table. For each document, descriptive data were also extracted, including information on the authors, year of publication, country, setting, participant characteristics, description of the intervention, description of the comparators, and other key findings related to the research question. One reviewer (FC or BN) extracted descriptive and outcome data, while a second reviewer (SP or FC) was responsible for verifying the data extraction for accuracy.

### Quality Assessment

The QUADAS-2 tool was used to assess the quality and risk of bias of each document retained for data extraction to critically appraise the validity of DBS testing (Whiting et al., 2006). One reviewer (FC or BN) assessed the quality of each document using the QUADAS-2 tool available from the QUADAS website (www.quadas.org), and a second reviewer (SP or FC) verified the assessment. A third reviewer, MB, resolved disagreements.

### Data Analysis

Data extracted from selected documents were synthesized through a narrative synthesis approach (Mays, Pope, & Popay, 2005). A narrative approach to synthesis was chosen because we had anticipated significant heterogeneity among documents in terms of context, patient populations, and index tests. Synthesized findings were compiled in tabulated form organized by STBBI, population, sample size, index test, and collection method. In addition, inductive thematic analysis was conducted in order to identify key themes and relationships in included studies.

## Results

We identified 7,631 abstracts in database searches. After 925 duplicates were removed, 6,706 titles and abstracts were screened, and 6,139 excluded. 567 reports were assessed for eligibility, with 397 further excluded. Of these, 230 were excluded due to no available data on measures of validity, 52 were excluded based on the study intervention, and 31 did not have full-text reports available. A further 23 studies were excluded for utilizing specimens other than blood, while 22 reports involved ineligible populations, and 15 were conducted for the purposes of genotyping or drug resistance studies designating them ineligible for inclusion. 16 studies were deemed ineligible on the basis of being unoriginal research. Eight reports were excluded as they were written in languages other than English or French. A total of 168 studies met the inclusion criteria and were included in the review. A PRISMA flow diagram (Page et al., 2021) representing the study selection is shown in Figure 1.

**Figure 1.**
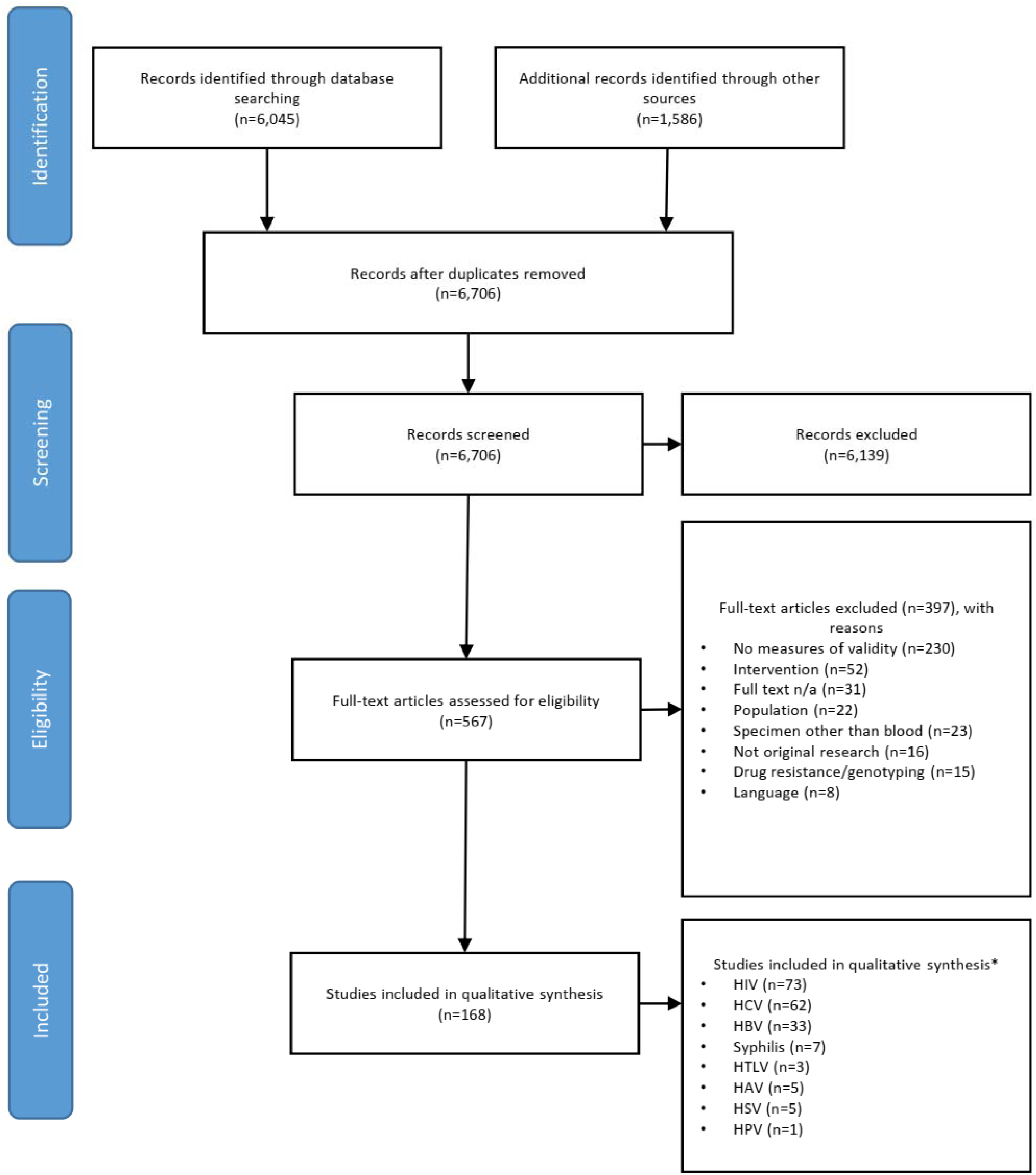
PRISMA flow diagram

### Study Characteristics

Characteristics of included studies are summarized in Supplementary File 2. A large proportion of included studies reported on the use of DBS for HIV testing (*n*=73; 43.5%). The remaining studies (*n=*115) reported on HCV (*n=*62; 36.9%), HBV (*n=*33; 19.6%), syphilis (*n=*7; 4.2%), HAV (*n=*5; 3.0%), HSV (*n=*4; 2.4%), HTLV (*n=*3; 1.8%), and HPV (*n=*1; 0.6%). No studies reported on chlamydia or gonorrhea. Several studies reported on more than one STBBI (e.g.,(Villar et al., 2011)), leading to a total of outcomes above the total number of studies included in this systematic review.

### STBBI Test Performance with DBS Specimens

Overall, included studies demonstrated high sensitivity and specificity for STBBI testing with DBS specimens (Fig. 2). HIV studies reported a wide range of values for sensitivity (9.0 to 100%) and specificity (4.0 to 100%). The majority of sensitivity measurements (*n*=137; 55.0%) were equal to or above 90.0%. Specificity measurements were also high, with most observations (*n*=172; 79.3%) above 90.0%. HCV studies reported sensitivity and specificity ranging from 36.0% to 100% and 85.7% to 100%, respectively. Most sensitivity (*n*=130; 77.4%) and specificity (*n*=116; 98.3%) measurements concerning HCV were equal to or above 90.0%. HBV studies reported sensitivity and specificity ranging from 40.0% to 100% and 2.5% to 100%,%, respectively. Approximately half of sensitivity measurements (*n*=34; 47.2%) and most specificity (*n*=60; 83.3%) measurements concerning HBV were equal to or above 90.0%. Studies involving syphilis reported high sensitivity and specificity, with ranges of 90.0% to 100% and 99.0% to 100%, respectively. HAV studies reported sensitivity and specificity ranging from 31.0% to 100% and 75.0% to 100%, respectively. Most sensitivity (*n*=4; 66.7%) and specificity (*n*=5; 83.3%) measurements concerning HAV were equal to or above 90.0%. HSV studies reported sensitivity and specificity ranging from 9.0% to 100% and 4.5% to 100%, respectively. Most sensitivity measurements (*n*=5; 55.6%) and almost half of specificity measurements (*n*=4; 44.4%) concerning HSV were equal to or above 90.0%. HTLV studies reported a range in sensitivity of 81.0% to 100%, while all included studies reported 100% specificity. The single HPV study reported an overall sensitivity and specificity of 98.0% and 92.0%, respectively. Approximately one-quarter of studies (*n*=47; 27.8%) reported LOD and/or LOQ values (Table 1). A meta-analysis was not undertaken due to the significant methodological heterogeneity among the included studies. This led us to identify several parameters that could influence index test performance.

**Figure 2.**
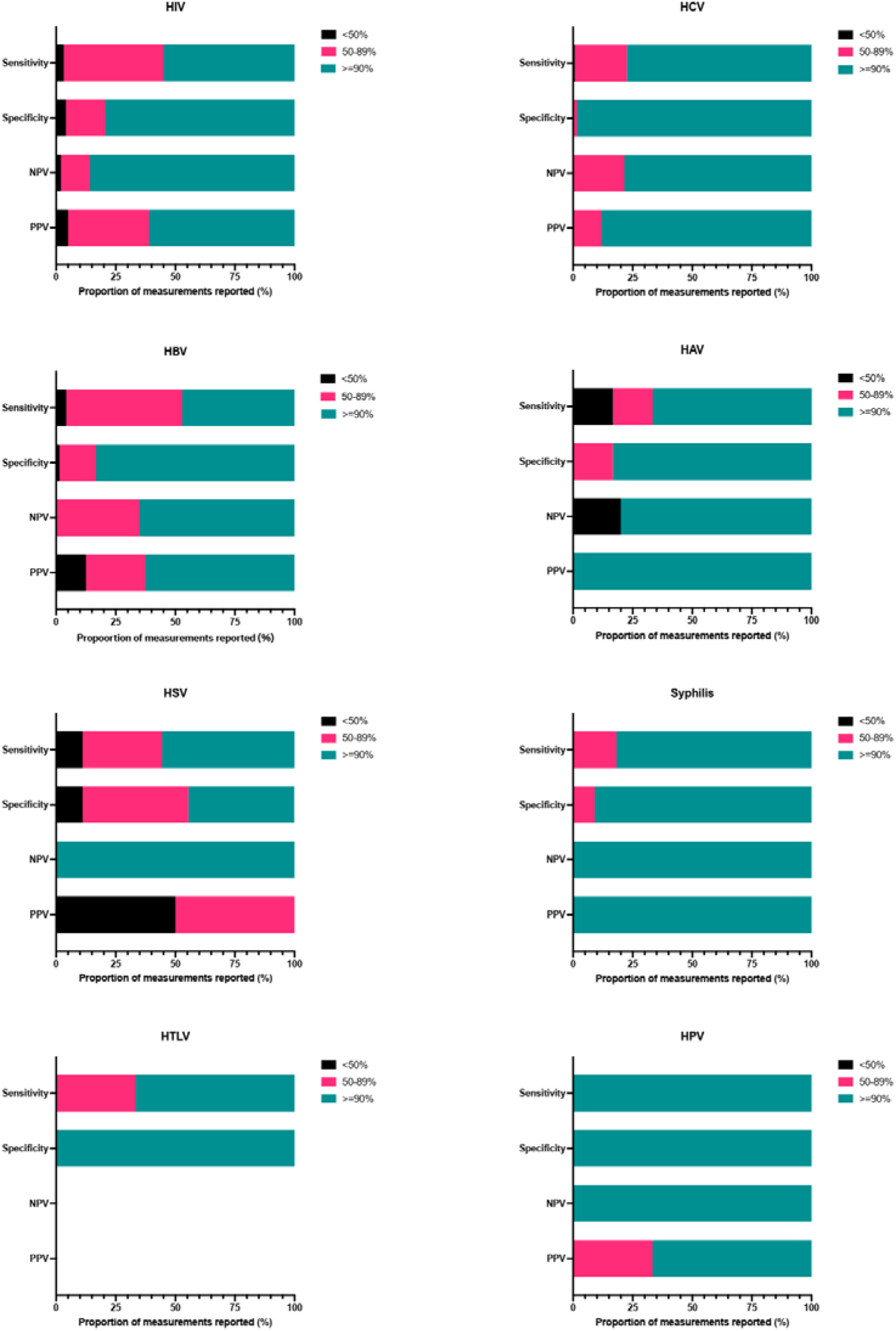
Proportion of reported performance measurements with <50%, 50-89%, or ≥90% sensitivity, specificity, negative predictive values (NPV), and positive predictive values (PPV)

**Table 1.** Studies reporting limit of detection and/or limit of quantification with DBS specimens

| Study | Index Test | STBBI | LOD | LOQ | Notes |
| --- | --- | --- | --- | --- | --- |
| (Aitken et al., 2013) | In-house RT-qPCR | HIV | 1,000 copies/mL | NR | DBS stored at ambient temperature (2 to 192 days) |
|  | In-house RT-qPCR | HIV | 1,000 copies/mL | NR | DBS stored at -20°C (45 to 112 days) |
|  | In-house RT-qPCR | HIV | 1,000 copies/mL | NR | DBS stored at -70°C (270 to 515 days) |
| (Patricia Alvarez et al., 2015) | COBAS AmpliPrep/TaqMan HIV-1 Quantitative Test v2.0 (Roche) | HIV | 400 copies/mL | NR | Mothers and infants (ART naïve) |
|  | COBAS AmpliPrep/TaqMan HIV-1 Quantitative Test v2.0 (Roche) | HIV | 400 copies/mL | NR | Mothers and infants (on ART) |
|  | COBAS AmpliPrep/TaqMan HIV-1 Quantitative Test v2.0 (Roche) | HIV | 400 copies/mL | NR | Mothers and infants (all) |
|  | Versant HIV-1 RNA 1.0 (Siemens) | HIV | ~880 copies/mL | NR | Mothers and infants (ART naïve) |
|  | Versant HIV-1 RNA 1.0 (Siemens) | HIV | ~880 copies/mL | NR | Mothers and infants (on ART) |
|  | Versant HIV-1 RNA 1.0 (Siemens) | HIV | ~880 copies/mL | NR | Mothers and infants (all) |
| (Andreotti et al., 2010) | COBAS TaqMan RT-qPCR (Roche) | HIV | 313 copies/mL | NR | Pregnant women (ART naïve) |
|  | COBAS TaqMan RT-qPCR (Roche) | HIV | 313 copies/mL | NR | Pregnant women (on ART) |
| (Choudhary et al., 2013) | In-house PCR | HIV | 1,300 CEM cells | NR | HIV-1 proviral test |
| (Erba et al., 2015) | m2000sp, m2000rt (Abbott) | HIV | 616 copies/mL | NR | (Erba et al., 2015) |
| (Fiscus, Brambilla, Grosso, Schock, & Cronin, 1998) | NASBA HIV-1 QT (Organon Teknika) | HIV | 1,000 copies/mL | NR |  |
| (Huang, Erickson, Mak, Salituro, & Abravaya, 2011) | RealTime HIV-1 Qualitative Assay (Abbott) | HIV | 2,469 copies/mL | NR | DBS prepared with material from the Virology Quality Assurance Laboratory (AIDS Clinical Trial Group) |
|  | RealTime HIV-1 Qualitative Assay (Abbott) | HIV | 3,085 copies/mL | NR | DBS prepared from the WHO 2<sup>nd</sup> International |
|  |  |  |  |  | Standard |
| (Kane et al., 2008) | NucliSENS HIV-1 EasyQ (bioMérieux) | HIV | 870 IU/mL | NR |  |
| (Marconi et al., 2009) | m2000sp, m2000rt (Abbott) | HIV | 200 – 1,000 copies/mL | NR |  |
| (Nugent et al., 2009) | Aptima HIV-1 RNA Qualitative Assay (Gen-Probe) | HIV | 2,384 copies/mL | NR |  |
| (Tang et al., 2017) | m2000 RealTime HIV-1 RNA (Abbott) | HIV | 839 copies/mL | 839 copies/mL |  |
| (Templer et al., 2016) | TaqMan HIV-1 v2.0 (Roche) | HIV | 221.8 copies/mL | NR |  |
|  | TaqMan HIV-1 v1.0 (Roche) | HIV | 1,090 copies/mL | NR |  |
|  | RealTime HIV-1 Qualitative (Abbott) | HIV | 2,500 copies/mL | NR |  |
| (van Deursen et al., 2010) | NucliSENS EasyQ HIV-1 v2.0 (bioMérieux) | HIV | 800 copies/mL | NR |  |
| (Viljoen et al., 2010) | Generic HIV Viral Load (Biocentric) | HIV | 3,100 copies/mL | NR | Durban (infants) |
|  | Generic HIV Viral Load (Biocentric) | HIV | 1,550 copies/mL | NR | Bobo-Dioulasso (mothers and infants) |
| (Zeh et al., 2017) | m2000 RealTime HIV-1 RNA (Abbott) | HIV | 1,222 copies/mL | NR |  |
| (Fajardo et al., 2014) | NucliSENS EasyQ HIV-1 v2.0 (bioMérieux) | HIV | NR | 100 copies/mL |  |
| (Mavedzenge et al., 2015) | NucliSENS EasyQ HIV-1 v2.0 (bioMérieux) | HIV | NR | 100 copies/mL |  |
| (Bennett et al., 2012) | In-house RT-qPCR | HCV | 250 IU/mL | NR |  |
| (Catlett et al., 2019) | Aptima HCV Quant DX (Hologic) | HCV | 13 IU/mL | 525 IU/mL |  |
| (De Crignis et al., 2010) | In-house RT-qPCR | HCV | 2,500 copies/mL | NR |  |
| (Mahajan, Choudhary, Kumar, & Gupta, 2018) | m2000rt HCV (Abbott) | HCV | 1,000 IU/mL | NR |  |
| (B. L. C. Marques et al., 2016) | In-house RT-qPCR | HCV | 58.5 copies/mL | NR |  |
| (Nguyen et al., 2018) | Generic HCV Assay (Biocentric) | HCV | 3,000 IU/mL | NR |  |
| (Vazquez-Moron et al., 2018) | In-house RT-qPCR | HCV | 5,000 copies/mL | NR |  |
| (Weber, Sahoo, Taylor, Shi, & Pinsky, 2019) | Aptima HCV Quant DX (Hologic) | HCV | 267 IU/mL | NR |  |
| (Parr et al., 2018) | m2000 RealTime HCV | HCV | 1,196 IU/mL | NR | 6 mm DBS |
|  | Viral Load (Abbott) |  |  |  | punches |
|  | m2000 RealTime HCV<br>Viral Load (Abbott) | HCV | 494 IU/mL | NR | 12 mm DBS<br>punches |
| (Santos et al.,<br>2012) | In-house RT-qPCR | HCV | 50 IU/mL | NR |  |
| (Shepherd, Baxter,<br>& Gunson, 2019) | m2000 RealTime HCV<br>Viral Load (Abbott) | HCV | 178 – 1,779<br>IU/mL | NR |  |
| (Solmone et al.,<br>2002) | In-house RT-qPCR | HCV | 960 IU/mL | NR |  |
| (Saludes et al.,<br>2018) | In-house RT-qPCR | HCV | 541 UI/mL | NR |  |
| (Stene-Johansen et<br>al., 2016) | In-house qPCR | HBV | 501.19 IU/mL | NR |  |
| (Jardi et al., 2004) | In-house qPCR | HBV | 2,000<br>copies/mL | NR |  |
| (Mohamed et al.,<br>2013) | Chemiluminescent<br>microparticle<br>immunoassay (Abbott) | HBV | 0.30 ± 0.08<br>IU/mL | NR | HBAg |
|  | Chemiluminescent<br>microparticle<br>immunoassay (Abbott) | HBV | 18.11 ± 6.05<br>IU/mL | NR | Anti-HBs |
|  | AmpliPrep/COBAS<br>TaqMan HBV Test v2.0<br>(Roche) | HBV | 914.1 ± 157.8<br>IU/mL | NR | HBV DNA |
| (Vinikoor et al.,<br>2015) | AmpliPrep/COBAS<br>TaqMan HBV v2.0<br>(Roche) | HBV | 1,000 IU/mL | NR |  |
| (Wilson P., 2018) | ARCHITECT<br>Qualitative HBsAg<br>Assay (Abbott) | HBV | 0.75 IU/mL | NR |  |
| (Lira et al., 2009) | HBV Monitor COBAS<br>Amplicor v1.5 (Roche) | HBV | 2,000<br>copies/mL | NR |  |
| (Zhang & Wang,<br>2010) | In-house PCR | HBV | 5 copies/mL | NR |  |
| (Jackson et al.,<br>2022) | RealTime HBV Viral<br>Load (Abbott) | HBV | 389 IU/mL | NR |  |
| (Roger et al., 2020) | Aptima HBV Quant<br>DX (Hologic) | HBV | 445 IU/mL | NR |  |
| (Shimakawa et al.,<br>2021) | LUMIPULSE CLEIA<br>(Fujirebio) | HBV | 19,115 IU/mL | NR | HBV core-related<br>antigen |
| (Bezerra et al.,<br>2022) | In-house qPCR | HBV | 852.5<br>copies/mL | NR |  |
| (Bezerra, Portilho,<br>Frota, & Villar,<br>2021) | In-house qPCR | HBV | 20 copies/mL | NR | High Pure Viral<br>Nucleic Acid Kit<br>(Roche) |
| (Bezerra et al.,<br>2021) | In-house PCR | HBV | 2,000<br>copies/mL | NR | QIAamp DNA<br>Blood Mini Kit<br>(Qiagen) |
|  | In-house PCR | HBV | 2,000<br>copies/mL | NR | Invisorb Spin<br>Blood Midi Kit<br>(Invitek) |
|  | In-house PCR | HBV | 20,000 | NR | DBS Genomic |
|  |  |  | copies/mL |  | DNA Isolation Kit (Norgen Biotek) |
| (Bargain, Heslan, Thibault, & Pronier, 2020) | GeneXpert HBV (Cepheid) | HBV | NR | 400 IU/mL |  |
| (Noda, Eizuru, Minamishima, Ikenoue, & Mori, 1993) | In-house PCR | HTLV | Single HUT102 cell | NR |  |

ART = antiretroviral therapy; DBS = dried blood spot; IU/mL = international units/mL; LOD = limit of detection; LOQ = limit of quantification; NR = not reported; STBBI = sexually transmitted and blood-borne infection

### Test Cut-Offs

Studies generally reported improved test performance by adjusting cut-off values – typically optimized for serum/plasma by manufacturers – for DBS specimens (Table 2). For example, García-Cisneros et al. (García-Cisneros et al., 2019) found that the IgG-G2 Human ELISA test (Human Diagnostics, Germany; HSV-2) performed better on DBS (specificity of 4.5% [95% CI=3%, 6.5%] versus 87.1% [95% CI=81.2%, 91.4%]) when using a cut-off value based on a receiver operating characteristic (ROC) curve. Villar et al. (Villar et al., 2011) also observed lower sensitivity (95.5% [95% CI=84.5%, 99.4%] versus 97.6% [95% CI=87.4%, 99.9%]; ETI-MAK-4 test, DiaSorin) and specificity (81.3% [95% CI=70.7%, 89.4%] versus 97.3% [95% CI=90.7%, 99.7%]; ETI-AB-AUK-3 test, DiaSorin) values when relying on manufacturer recommended cut-offs compared to cut-off values based on ROC curve analysis for the detection of HBV in DBS specimens. Ultimately, we observed that the choice of cut-off value (manufacturer recommended versus established in-house) influenced test performance when analyzing DBS specimens.

**Table 2.** Studies reporting test performance according to test cut-off values

| Study | STBBI | Index Test | Cut-Off | Sensitivity (95% CI) | Specificity (95% CI) | PPV (95% CI) | NPV (95% CI) | Notes |
| --- | --- | --- | --- | --- | --- | --- | --- | --- |
| (García-Cisneros et al., 2019) | HSV-2 | IgG-G2 Human (Human Diagnostics) | ≥1.15<sup>a</sup> | 99.7 (98.3-100.0) | 4.5 (3.0-6.5) | 37.3 (34.1-40.6) | 96.3 (81.0-99.9) |  |
|  |  |  | ≥4.61<sup>b</sup> | 90.4 (84.4-94.4) | 87.1 (81.2-91.4) | 84.7 (77.9-89.8) | 92.0 (86.7-95.4) |  |
| (Villar et al., 2011) | HBV | ETI-AB-COREK Plus (DiaSorin) | Mean of 3 calibrators x 0.3<sup>a</sup> | 89.2 (79.8-95.2) | 97.5 (91.4-99.7) | 97.1 (89.8-99.6) | 90.8 (82.7-96.0) |  |
|  |  |  | ≤0.951<sup>b</sup> | 100.0 (95.1-100.0) | 2.5 (0.3-8.6) | 48.4 (40.2-56.6) | 100.0 (15.8-100.0) |  |
|  |  |  | ≤0.261<sup>b</sup> | 90.5 (81.5-96.1) | 92.6 (84.6-97.2) | 91.8 (83.0-96.9) | 91.5 (83.2-96.5) |  |
|  |  | ETI-AB-AUK-3 (DiaSorin) | Mean of kit calibrator 1<sup>a</sup> | 83.1 (71.0-91.6) | 81.3 (70.7-89.4) | 77.8 (65.5-87.3) | 85.9 (75.6-93.0) |  |
|  |  |  | >0.119<sup>b</sup> | 66.1 (52.6-77.6) | 98.7 (92.8-100.0) | 97.5 (86.8-99.9) | 78.7 (69.1-86.5) |  |
|  |  |  | >0.095<sup>b</sup> | 78.0 (65.3-87.7) | 97.3 (90.7-99.7) | 95.8 (85.8-99.5) | 84.9 (75.5-91.7) |  |
|  |  | ETI-MAK-4 (DiaSorin) | Mean of negative control + 0.03<sup>a</sup> | 95.5 (84.5-99.4) | 70.8 (60.2-80.0) | 61.8 (49.2-73.3) | 96.9 (89.3-99.6) |  |
|  |  |  | >0.100<sup>b</sup> | 93.2 | 98.9 | 97.6 | 96.7 |  |
|  |  |  |  | (81.3-98.6) | (93.9-100.0) | (87.4-99.9) | (90.7-99.3) |  |
|  |  |  | >0.115<sup>b</sup> | 97.6<br>(87.4-99.9) | 96.7<br>(90.7-99.3) | 93.2<br>(81.3-98.6) | 98.9<br>(93.9-100.0) |  |
| (Catlett et al., 2019) |  | Aptima HCV Quant Dx (Hologic) | Detected | 96.4<br>(89.8-99.3) | 95.8<br>(78.9-99.9) | NR | NR |  |
|  |  |  | ≥15 IU/mL | 95.1<br>(88.0-98.7) | 96.0<br>(79.7-99.9) | NR | NR |  |
|  |  |  | ≥1,000 IU/mL | 100.0<br>(95.3-100.0) | 100.0<br>(88.4-100.0) | NR | NR |  |
| (Brandão et al., 2013) | HCV | Monolisa HCV AgAb ULTRA EIA (Bio-Rad) | Not specified<sup>a</sup> | 95.0<br>(83.1-99.4) | 100.0<br>(98.9-100.0) | 100.0<br>(90.8-100.0) | 99.4<br>(97.9-99.9) |  |
|  |  |  | 0.108<sup>b</sup> | 97.5<br>(86.8-99.9) | 97.2<br>(94.9-98.6) | 79.6<br>(65.7-89.8) | 99.7<br>(98.4-99.0) |  |
|  |  |  | 0.287<sup>b</sup> | 97.5<br>(86.8-99.9) | 99.7<br>(98.4-100.0) | 100.0<br>(91.0-100.0) | 100.0<br>(99.0-100.0) |  |
|  |  | Murex HCV AgAb Combination EIA (DiaSorin) | Not specified<sup>a</sup> | 82.5<br>(67.2-92.7) | 98.0<br>(95.9-99.2) | 82.5<br>(67.2-92.7) | 98.0<br>(95.9-99.2) |  |
|  |  |  | 0.514<sup>b</sup> | 80.0<br>(64.3-90.9) | 99.4<br>(97.9-99.9) | 94.1<br>(80.3-99.3) | 97.7<br>(95.6-99.0) |  |
|  |  |  | 0.239<sup>b</sup> | 97.5<br>(86.8-99.9) | 96.0<br>(93.3-97.8) | 73.6<br>(93.3-97.8) | 100<br>(98.3-100.0) |  |
| (Erba et al., 2015) | HIV | m2000SP, m2000RT (Abbott) | ≥40 copies/mL | 74.0<br>(66.9-80.3) | 99.0<br>(94.5-99.8) | 99.2<br>(95.8-99.9) | 68.3<br>(60.0-75.8) |  |
|  |  |  | ≥1,000 copies/mL | 94.2<br>(88.9-97.5) | 98.6<br>(94.9-99.8) | 98.5<br>(94.6-99.8) | 94.5<br>(89.4-97.6) |  |
| (Fajardo et al., 2014) | HIV | NucliSENS EasyQ HIV-1 v2.0 (bioMérieux) | ≥1,000 copies/mL | 88.7<br>(81.1-94.0) | 97.8<br>(96.1-98.9) | NR | NR | DBS prepared from capillary blood (finger pokes) |
|  |  |  | ≥3,000 copies/mL | 84.9 | 99.8 | NR | NR | DBS prepared |
|  |  |  |  | (76.0-91.5) | (98.9-100.0) |  |  | from capillary blood (finger pokes) |
|  |  |  | ≥5,000 copies/mL | 83.0 (73.4-90.1) | 100.0 (99.3-100.0) | NR | NR | DBS prepared from capillary blood (finger pokes) |
|  |  |  | ≥1,000 copies/mL | 91.4 (84.4-96.0) | 97.2 (95.4-98.5) | NR | NR | DBS prepared from venous blood |
|  |  |  | ≥3,000 copies/mL | 90.0 (82.2-95.4) | 99.4 (98.3-99.9) | NR | NR | DBS prepared from venous blood |
|  |  |  | ≥5,000 copies/mL | 88.5 (79.9-94.3) | 99.8 (98.9-100.0) | NR | NR | DBS prepared from venous blood |
| (Guichet et al., 2018) | HIV | In-house RT-qPCR | ≥1,000 copies/mL | 90.0 (83.0-96.0) | 49.0 (37.0-61.0) | NR | NR | NucliSENS (bioMérieux) |
|  |  |  | ≥5,000 copies/mL | 74.0 (62-86.0) | 77.0 (69.0-85.0) | NR | NR | NucliSENS (bioMérieux) |
|  |  |  | ≥1,000 copies/mL | 80.0 (71.0-89.0) | 59.0 (47.0-71.0) | NR | NR | m2000SP (Abbott) |
|  |  |  | ≥5,000 copies/mL | 79.0 (68.0-90.0) | 68.0 (59.0-72.0) | NR | NR | m2000SP (Abbott) |
|  |  |  | ≥1,000 copies/mL | 80.0 (71.0-89.0) | 100.0 (100.0-100.0) | NR | NR | NucliSENS (bioMérieux), Turbo DNase-Free (Ambion) |
|  |  |  | ≥5,000 copies/mL | 23.0 (12.0-34.0) | 100.0 (100.0-100.0) | NR | NR | NucliSENS (bioMérieux), Turbo DNase-Free (Ambion) |
|  |  |  | ≥1,000 copies/mL | 27.0 (17.0-37.0) | 81.0 (70.0-90.0) | NR | NR | NucliSENS (bioMérieux), HL-dsDNase (TATAA Biocenter AB) |
|  |  |  | ≥5,000 copies/mL | 51.0 (37.0-65.0) | 92.0 (86.0-97.0) | NR | NR | NucliSENS (bioMérieux), HL-dsDNase (TATAA |
|  |  |  |  |  |  |  |  | Biocenter AB) |
|  |  |  | ≥1,000 copies/mL | 60.0 (49.0-71.0) | 82.0 (73.0-91.0) | NR | NR | NucliSENS (bioMérieux), FVE protocol |
|  |  |  | ≥5,000 copies/mL | 49.0 (35.0-62.0) | 99.0 (97.0-100.0) | NR | NR | NucliSENS (bioMérieux), FVE protocol |
|  |  |  | ≥1,000 copies/mL | 83.0 (75.0-91.0) | 58.0 (46.0-70.0) | NR | NR | m2000SP, m2000RT (Abbott) |
|  |  |  | ≥5,000 copies/mL | 75.0 (63.0-87.0) | 89.0 (83.0-95.0) | NR | NR | m2000SP, m2000RT (Abbott) |
| (Halfon et al., 2012) | HBV | HBV COBAS TaqMan (Roche) | ≥1,400 IU/mL | 98.0 (95.0-100.0) | 100.0 (100.0-100.0) | NR | NR |  |
|  |  |  | ≥2,000 IU/mL | 91.0 (85.0-97.0) | 100.0 (100.0-100.0) | NR | NR |  |
| (Hobbs et al., 2017) | HSV-2 | Kalon HSV-2 ELISA (Kalon Biological Ltd) | >1.1<sup>a</sup> | 98.8 (92.7-99.9) | 98.9 (93.4-99.9) | NR | NR |  |
|  |  |  | >1.5<sup>b</sup> | 98.8 (93.6-99.8) | 98.9 (94.2-99.8) | NR | NR |  |
| (Judd et al., 2003) | HCV | Ortho HCV 3.0 (Bio-Rad) | 0.100<sup>a</sup> | 99.2 | 100.0 |  |  |  |
|  |  |  | Negative mean + 1 SD<sup>b</sup> | 100.0 | 92.4 | NR | NR |  |
|  |  |  | Negative mean + 2 SD<sup>b</sup> | 100.0 | 96.9 | NR | NR |  |
|  |  |  | Negative mean + 3 SD<sup>b</sup> | 99.6 | 99.7 | NR | NR |  |
|  |  |  | Negative mean + 4 SD<sup>b</sup> | 99.6 | 99.7 | NR | NR |  |
|  |  |  | Negative mean + 5 SD<sup>b</sup> | 99.6 | 100.0 | NR | NR |  |
|  |  |  | Negative mean + 6 SD<sup>b</sup> | 99.2 | 100.0 | NR | NR |  |
| (Makadzang et al., 2017) | HIV | COBAS AmpliPrep/COBAS TaqMan (Roche) | Detected | 77.0 (69.4-85.5) | 100.0 (97.1-100.0) | 100.0 (96.8-100.0) | 78.5 (71.2-84.6) |  |
|  |  |  | ≥1,000<br>copies/mL | 92.7<br>(88.6-96.6) | 100.0<br>(97.6-100.0) | 100.0<br>(96.8-100.0) | 94.3<br>(89.5-97.4) |  |
|  |  |  | ≥5,000<br>copies/mL | 70.9<br>(61.8-79.0) | 100.0<br>(97.6-100.0) | 100.0<br>(95.7-100.0) | 82.0<br>(75.8-87.2) |  |
| (B. L. Marques et al., 2012) | HCV | ETI-AB-HCVK-4 (DiaSorin) | 0.592<sup>a</sup> | 88.9<br>(76.0-96.3) | 96.1<br>(93.6-97.9) | 74.1<br>(60.4-85.0) | 98.6<br>(96.7-99.5) |  |
|  |  |  | 0.648<sup>b</sup> | 88.9<br>(76.0-96.3) | 96.4<br>(93.9-98.1) | 75.5<br>(61.7-86.2) | 98.6<br>(96.7-99.5) |  |
|  |  |  | 1.345<sup>b</sup> | 88.9<br>(76.0-96.3) | 98.6<br>(96.8-99.6) | 88.9<br>(76.0-96.3) | 98.6<br>(96.8-99.6) |  |
|  |  | HCV Ab (Radim) | 0.347<sup>a</sup> | 97.5<br>(86.8-99.9) | 99.5<br>(98.1-99.9) | 95.1<br>(83.5-99.4) | 99.7<br>(98.5-100.0) |  |
|  |  |  | 0.351<sup>b</sup> | 97.5<br>(86.8-99.9) | 99.5<br>(98.1-99.9) | 95.1<br>(83.5-99.4) | 99.7<br>(98.5-100.0) |  |
|  |  |  | 0.284<sup>b</sup> | 97.5<br>(86.8-99.9) | 99.2<br>(97.7-99.8) | 92.9<br>(80.5-98.5) | 99.7<br>(98.5-100.0) |  |
| (McCarron et al., 1999) | HCV | Monolisa anti-HCV (Sanofi Pasteur) | >0.99<sup>b</sup> | 100.0 | 87.5 | NR | NR |  |
|  |  |  | >1.99<sup>b</sup> | 97.2 | 100.0 | NR | NR |  |
| (Neogi et al., 2012) | HIV | m2000RT, m2000SP (Abbott) | 2.17 to 3<br>log<sup>10</sup><br>copies/mL | 50.0 | 100.0 | NR | NR |  |
|  |  |  | >3 to 3.7<br>log<sup>10</sup><br>copies/mL | 90.0 | 100.0 | NR | NR |  |
|  |  |  | >3.7 log<sup>10</sup><br>copies/mL | 100.0 | 100.0 | NR | NR |  |
| (Nugent et al., 2009) | HIV | Aptima HIV-1 RNA Qualitative Assay (Gen-Probe) | ≥500<br>copies/mL | 65.0 | NR | NR | NR |  |
|  |  |  | ≥2,500<br>copies/mL | 92.0 | NR | NR | NR |  |
|  |  |  | ≥5,000<br>copies/mL | 100 | NR | NR | NR |  |
|  |  |  | ≥10,000<br>copies/mL | 100 | NR | NR | NR |  |
| (Pannus et al., 2013) | HIV | NucliSENS EasyQ v2.0 (bioMérieux) | ≥1,000 copies/mL | 78.6 (59.0-91.7) | 100.0 (98.9-100.0) | 100.0 (84.6-100.0) | 98.2 (96.1-99.3) | DBS prepared from capillary blood (finger pokes) |
|  |  |  | ≥1,000 copies/mL | 89.3 (71.8-97.7) | 99.7 (98.3-100.0) | 96.2 (80.4-99.9) | 99.1 (97.3-99.8) | DBS prepared from venous blood |
|  |  |  | ≥5,000 copies/mL | 69.6 (47.1-86.8) | 100.0 (89.9-100.0) | 100.0 (79.4-100.0) | 97.9 (95.7-99.2) | DBS prepared from capillary blood (finger pokes) |
|  |  |  | ≥5,000 copies/mL | 60.9 (38.5-80.3) | 100.0 (98.9-100.0) | 100.0 (76.8-100.0) | 97.3 (95.0-98.8) | DBS prepared from venous blood) |
| (Pirillo et al., 2011) | HIV | VERSANT HIV-1 RNA 1.0 (Siemens) | ≥37 copies/mL | 88.2 (79.4-93.6) | 69.2 (42.0-87.4) | 94.9 (90.1-99.8) | 47.4 (24.9-69.8) |  |
|  |  |  | ≥5,000 copies/mL | 85.1 (76.5-88.6) | 96.1 (88.2-99.3) | 95.2 (85.6-99.1) | 87.5 (80.3-90.4) |  |
| (Pollack et al., 2018) | HIV | COBAS AmpliPrep/COBAS TaqMan (Roche) | ≥1,000 copies/mL | 98.8 (93.3-100.0) | 74.3 (70.8-77.5) | 31.5 (25.8-37.6) | 99.8 (98.9-100.0) | Guanidinium pre-extraction |
|  |  |  | ≥5,000 copies/mL | 92.4 (84.2-97.2) | 97.9 (96.6-98.9) | 83.9 (74.5-90.9) | 99.1 (98.1-99.7) | Guanidinium pre-extraction |
|  |  |  | ≥1,000 copies/mL | 95.1 (87.8-98.6) | 98.8 (97.7-99.5) | 90.6 (82.3-95.8) | 99.4 (98.5-99.8) | Correction factor applied (0.3 log<sup>10</sup> copies/mL) |
|  |  |  | ≥5,000 copies/mL | 96.3 (89.6-99.2) | 98.2 (96.9-99.1) | 86.7 (77.9-92.9) | 99.6 (98.7-99.9) | Correction factor applied (0.3 log<sup>10</sup> copies/mL) |
|  |  |  | ≥1,000 copies/mL | 65.8 (54.3-76.1) | 100.0 (99.5-100.0) | 100.0 (93.2-100.0) | 96.2 (94.5-97.5) | Correction factor applied (0.7 log<sup>10</sup> copies/mL) |
|  |  |  | ≥5,000 copies/mL | 84.8 (75.0-91.9) | 99.7 (98.9-100.0) | 97.1 (89.9-99.6) | 98.3 (97.0-99.1) | Correction factor applied (0.7 |

|  |  |  |  |  |  |  |  | log<sup>10</sup><br>copies/mL) |
| --- | --- | --- | --- | --- | --- | --- | --- | --- |
| (Rutstein et al., 2014) | HIV | m2000RT, m2000SP (Abbott) | ≥1,000 copies/mL | 100.0 | 97.1 | 75.0 | 100.0 | DBS prepared from venous blood |
|  |  |  | ≥1,000 copies/mL | 100.0 | 94.9 | 63.2 | 100.0 | DBS prepared from capillary blood (finger pokes) |
|  |  |  | ≥5,000 copies/mL | 100.0 | 98.6 | 83.3 | 100.0 | DBS prepared from venous blood |
|  |  |  | ≥5,000 copies/mL | 100.0 | 97.8 | 76.9 | 100.0 | DBS prepared from capillary blood (finger pokes) |
| (Sawadogo et al., 2014) | HIV | COBAS AmpliPrep/COBAS TaqMan (Roche) | ≥1,000 copies/mL | 99.0 (97.0-100.0) | 26.0 (22.0-29.0) | 29.0 (26.0-33.0) | 99.0 (96.0-100.0) |  |
|  |  |  | ≥5,000 copies/mL | 99.0 (96.0-100.0) | 55.0 (51.0-59.0) | 33.0 (29.0-37.0) | 100.0 (98.0-100.0) |  |
| (Schmitz et al., 2017) | HIV | COBAS AmpliPrep/COBAS TaqMan (Roche) | ≥1,000 copies/mL | 90.1 (85.7-93.6) | 93.1 (90.6-95.2) | NR | NR | DBS prepared from venous blood |
|  |  | COBAS AmpliPrep/COBAS TaqMan (Roche) | ≥1,000 copies/mL | 90.3 (85.7-93.7) | 94.9 (92.6-96.6) | NR | NR | DBS prepared from capillary blood (microtainer) |
|  |  | COBAS AmpliPrep/COBAS TaqMan (Roche) | ≥1,000 copies/mL | 88.1 (83.3-92.0) | 94.5 (92.1-96.3) | NR | NR | DBS prepared from capillary blood (finger pokes) |
|  |  | RealTime HIV-1 (Abbott) | ≥1,000 copies/mL | 94.4 (90.6-97.0) | 33.0 (28.9-37.3) | NR | NR | DBS prepared from venous blood |
|  |  | COBAS AmpliPrep/COBAS TaqMan (Roche) | ≥3,000 copies/mL | 85.2 (80.1-89.4) | 98.0 (96.4-99.1) | NR | NR | DBS prepared |
|  |  | S TaqMan (Roche) |  |  |  |  |  | from venous blood |
|  |  | COBAS AmpliPrep/COBAS TaqMan (Roche) | ≥3,000 copies/mL | 88.1 (83.3-92.0) | 97.6 (95.9-98.8) | NR | NR | DBS prepared from capillary blood (microtainer) |
|  |  | COBAS AmpliPrep/COBAS TaqMan (Roche) | ≥3,000 copies/mL | 85.2 (80.0-89.4) | 97.8 (96.2-98.9) | NR | NR | DBS prepared from capillary blood (finger pokes) |
|  |  | RealTime HIV-1 (Abbott) | ≥3,000 copies/mL | 88.4 (83.5-92.2) | 60.9 (56.6-65.2) | NR | NR | DBS prepared from venous blood |
|  |  | COBAS AmpliPrep/COBAS TaqMan (Roche) | ≥5,000 copies/mL | 83.1 (77.8-87.6) | 98.4 (96.9-99.3) | NR | NR | DBS prepared from venous blood |
|  |  | COBAS AmpliPrep/COBAS TaqMan (Roche) | ≥5,000 copies/mL | 83.9 (78.6-88.3) | 98.6 (97.2-99.4) | NR | NR | DBS prepared from capillary blood (microtainer) |
|  |  | COBAS AmpliPrep/COBAS TaqMan (Roche) | ≥5,000 copies/mL | 82.2 (76.7-86.9) | 98.8 (97.4-99.6) | NR | NR | DBS prepared from capillary blood (finger pokes) |
|  |  | RealTime HIV-1 (Abbott) | ≥5,000 copies/mL | 79.7 (74.0-84.7) | 77.0 (73.1-80.5) | NR | NR | DBS prepared from venous blood |
| (Taieb et al., 2018) | HIV | m2000SP, m2000RT (Abbott) | Detected | 95.8 (88.1-99.1) | 63.6 (54.8-71.8) | NR | NR |  |
|  |  |  | ≥839 copies/mL | 91.5 (82.5-96.8) | 95.5 (90.3-98.3) | NR | NR |  |
|  |  |  | ≥1,000 copies/mL | 90.1 (80.7-95.9) | 96.2 (91.4-98.8) | NR | NR |  |
|  |  |  | ≥3,000 copies/mL | 84.5 (74.0-92.0) | 99.2 (95.9-100.0) | NR | NR |  |
|  |  |  | ≥5,000 copies/mL | 80.3 (69.1-88.8) | 100.0 (97.2-100.0) | NR | NR |  |
| (Taieb et al., | HIV | COBAS | ≥1,000 | 54.9 | 100.0 | NR | NR |  |
| 2016) |  | AmpliPrep/COBAS TaqMan HIV-1 v2.0 (Roche) | copies/mL | (40.3-68.9) | (97.5-100.0) |  |  |  |
|  |  |  | ≥3,000 copies/mL | 45.2 (29.8-61.3) | 100.0 (97.7-100.0) | NR | NR |  |
|  |  |  | ≥5,000 copies/mL | 47.5 (31.5-63.8) | 100.0 (97.7-100.0) | NR | NR |  |
|  |  | Real-Time HIV-1 (Abbott) | ≥1,000 copies/mL | 93.3 (81.7-98.6) | 94.8 (90.0-97.7) | NR | NR |  |
|  |  |  | ≥3,000 copies/mL | 97.4 (86.2-99.9) | 96.3 (92.0-98.6) | NR | NR |  |
|  |  |  | ≥5,000 copies/mL | 100 (90.5-100.0) | 98.8 (95.6-99.8) | NR | NR |  |
| (Vidya et al., 2012) |  | m2000RT (Abbott) | ≤1,000 copies/mL | 62.0 | NR | NR | NR |  |
|  |  |  | 1,000-3,000 copies/mL | 88.0 | NR | NR | NR |  |
|  |  |  | >3,000 copies/mL | 100.0 | NR | NR | NR |  |
| (Zeh et al., 2017) | HIV | COBAS AmpliPrep/COBAS TaqMan (Roche) | ≥400 copies/mL | 100.0 (97.6-100.0) | 4.0 (0.5-13.7) | 75.8 (69.2-81.6) | 100.0 (15.8-100.0) |  |
|  |  |  | ≥1,000 copies/mL | 100.0 | 17.3 | NR | NR |  |
|  |  |  | ≥2,000 copies/mL | 98.0 | 36.5 | NR | NR |  |
|  |  |  | ≥3,000 copies/mL | 97.3 | 54.0 | NR | NR |  |
|  |  |  | ≥4,000 copies/mL | 96.0 | 82.7 | NR | NR |  |
|  |  |  | ≥5,000 copies/mL | 95.3 | 84.6 | NR | NR |  |
|  |  | m2000RT (Abbott) | ≥550 copies/mL | 93.9 (88.8-97.2) | 88.0 (82.2-92.4) | 100.0 (97.4-100.0) | 85.3 (73.8-93.0) |  |
|  |  |  | ≥1,000 copies/mL | 96.6 | 90.4 | NR | NR |  |
|  |  |  | ≥2,000 copies/mL | 95.3 | 94.2 | NR | NR |  |
|  |  |  | ≥3,000 copies/mL | 94.6 | 98.1 | NR | NR |  |
|  |  |  | ≥4,000 copies/mL | 93.2 | 98.1 | NR | NR |  |
|  |  |  | ≥5,000<br>copies/mL | 93.0 | 98.1 | NR | NR |  |
| (Biondi et al.,<br>2019) | HCV | ARCHITECT Core<br>Antigen (Abbott) | >3 fmol/L | 94.1<br>(88.5-99.7) | NR | NR | NR | DBS stored<br>at -80°C |
|  |  |  | >10<br>fmol/L | 85.3<br>(76.4-94.2) | NR | NR | NR | DBS stored<br>at -80°C |
|  |  |  | >3 fmol/L | 94.1<br>(88.5-99.7) | NR | NR | NR | DBS stored<br>at 4°C |
|  |  |  | >10<br>fmol/L | 85.3<br>(76.4-94.2) | NR | NR | NR | DBS stored<br>at 4°C |
|  |  |  | >3 fmol/L | 91.2<br>(84.3-98.1) | NR | NR | NR | DBS stored<br>at ambient<br>°C |
|  |  |  | >10<br>fmol/L | 80.9<br>(70.8-91.0) | NR | NR | NR | DBS stored<br>at ambient<br>°C |
|  |  |  | >3 fmol/L | 92.7<br>(86.4-98.9) | NR | NR | NR | DBS stored<br>at 37°C |
|  |  |  | >10<br>fmol/L | 80.9<br>(70.8-91.0) | NR | NR | NR | DBS stored<br>at 37°C |
|  |  |  | >3 fmol/L | 92.7<br>(86.4-98.9) | NR | NR | NR | DBS stored<br>at 37°C<br>followed by<br>4°C |
|  |  |  | >10<br>fmol/L | 85.3<br>(76.4-94.2) | NR | NR | NR | DBS stored<br>at 37°C<br>followed by<br>4°C |
| (Catlett et al.,<br>2019) | HCV | Aptima HCV<br>Quant Dx<br>(Hologic) | ≥12<br>IU/mL | 90.7<br>(80.0-97.0) | 100.0<br>(97.0-100.0) | NR | NR |  |
|  |  |  | ≥1,000<br>IU/mL | 92.5<br>(82.0-98.0) | 100.0<br>(97.0-100.0) | NR | NR |  |
|  |  |  | ≥3,000<br>IU/mL | 92.5<br>(92.0-98.0) | 100.0<br>(97.0-100.0) | NR | NR |  |
| (Saludes et<br>al., 2019) | HCV | In-house RT-qPCR | ~12<br>IU/mL | 88.3<br>(83.0-92.0) | NR | NR | NR |  |
|  |  |  | ≥12<br>IU/mL | 90.1<br>(85.0-93.5) | NR | NR | NR |  |
|  |  |  | ≥1,000<br>copies/mL | 96.1<br>(92.1-98.1) | NR | NR | NR |  |
|  |  |  | ≥3,000<br>copies/mL | 97.2 | NR | NR | NR |  |
|  |  |  |  | (93.5-98.8) |  |  |  |  |
|  |  |  | ≥50,000<br>copies/mL | 100.0<br>(97.6-100.0) | NR | NR | NR |  |
| (Saludes et al., 2020) | HCV | Xpert HCV VL<br>Fingerstick<br>(Cepheid) | ~10<br>IU/mL | 93.7<br>(84.8-97.5) | 100.0<br>(90.1-100.0) | NR | NR |  |
|  |  |  | ≥10<br>IU/mL | 96.7<br>(88.8-99.1) | 100.0<br>(90.1-100.0) | NR | NR |  |
|  |  |  | ≥1,000<br>IU/mL | 98.3<br>(91.1-99.7) | 100.0<br>(90.1-100.0) | NR | NR |  |
|  |  |  | ≥3,000<br>IU/mL | 98.3<br>(91.0-99.7) | 100.0<br>(90.1-100.0) | NR | NR |  |
| (Tola et al., 2021) | HIV | m2000 RealTime<br>HIV-1 (Abbott) | ≥1,000<br>copies/mL | 85.2<br>(81.1-89.3) | 91.5<br>(88.7-94.3) | NR | NR | DBS<br>prepared<br>from<br>capillary<br>blood<br>(microtainer) |
|  |  |  | ≥3,000<br>copies/mL | 72.5<br>(67.3–77.7) | 96.9<br>(95.2–98.6) | NR | NR | DBS<br>prepared<br>from<br>capillary<br>blood<br>(microtainer) |
|  |  |  | ≥5,000<br>copies/mL | 65.8<br>(60.3–71.4) | 97.2<br>(95.5–98.8) | NR | NR | DBS<br>prepared<br>from<br>capillary<br>blood<br>(microtainer) |
|  |  |  | ≥1,000<br>copies/mL | 85.6<br>(81.5–89.6) | 90.9<br>(88.1–93.8) | NR | NR | DBS<br>prepared<br>from<br>capillary<br>blood (finger<br>pokes) |
|  |  |  | ≥3,000<br>copies/mL | 71.5<br>(66.2–76.7) | 96.6<br>(94.9–98.4) | NR | NR | DBS<br>prepared<br>from<br>capillary<br>blood (finger<br>pokes) |
|  |  |  | ≥5,000 | 62.7 | 97.4 | NR | NR | DBS |
|  |  |  | copies/mL | (57.1–68.3) | (95.9–99.0) |  |  | prepared from capillary blood (finger pokes) |
|  |  |  | ≥1,000 copies/mL | 89.1 (85.5–92.7) | 86.6 (83.2–90.0) | NR | NR | DBS prepared from venous blood |
|  |  |  | ≥3,000 copies/mL | 76.4 (71.5–81.3) | 96.1 (94.2–98.1) | NR | NR | DBS prepared from venous blood |
|  |  |  | ≥5,000 copies/mL | 67.2 (61.8–72.7) | 96.6 (95.2–98.6) | NR | NR | DBS prepared from venous blood |

<sup>a</sup>Manufacturer recommended
<sup>b</sup>Established in-house
NPV=negative predictive value; NR=not reported; PPV=positive predictive value; STBBI=sexually transmitted and blood-borne infection;

### DBS Specimen Preparation

DBS specimens prepared from venous blood instead of capillary blood (i.e., finger pokes) appear to produce higher sensitivity and specificity values (Table 3). This was particularly evident with regards to HIV. Multiple studies reported better HIV serological and nucleic acid test performance when analyzing DBS specimens prepared from venous blood (Fajardo et al., 2014; Mwau et al., 2021; Rutstein et al., 2014). A few papers examined test performance between DBS specimens prepared from venous and capillary blood for HCV testing, with the majority of those finding similar or greater sensitivity when using DBS specimens prepared with venous blood. Prinsenberg et al. (Prinsenberg et al., 2020) reported a small advantage in sensitivity (96.4% [95% CI=81.7%, 99.9%] versus 95.7% [78.1%, 99.9]) when using venous blood to prepare DBS specimens, while Tran et al. (Tran et al., 2020) and Vetter et al. (Vetter et al., 2021) both reported nearly identical test performance with DBS prepared from venous and capillary blood (Table 3). In general, how DBS are prepared (venous versus capillary blood) could influence test performance, with DBS prepared from venous blood contributing to better test performance.

**Table 3.**
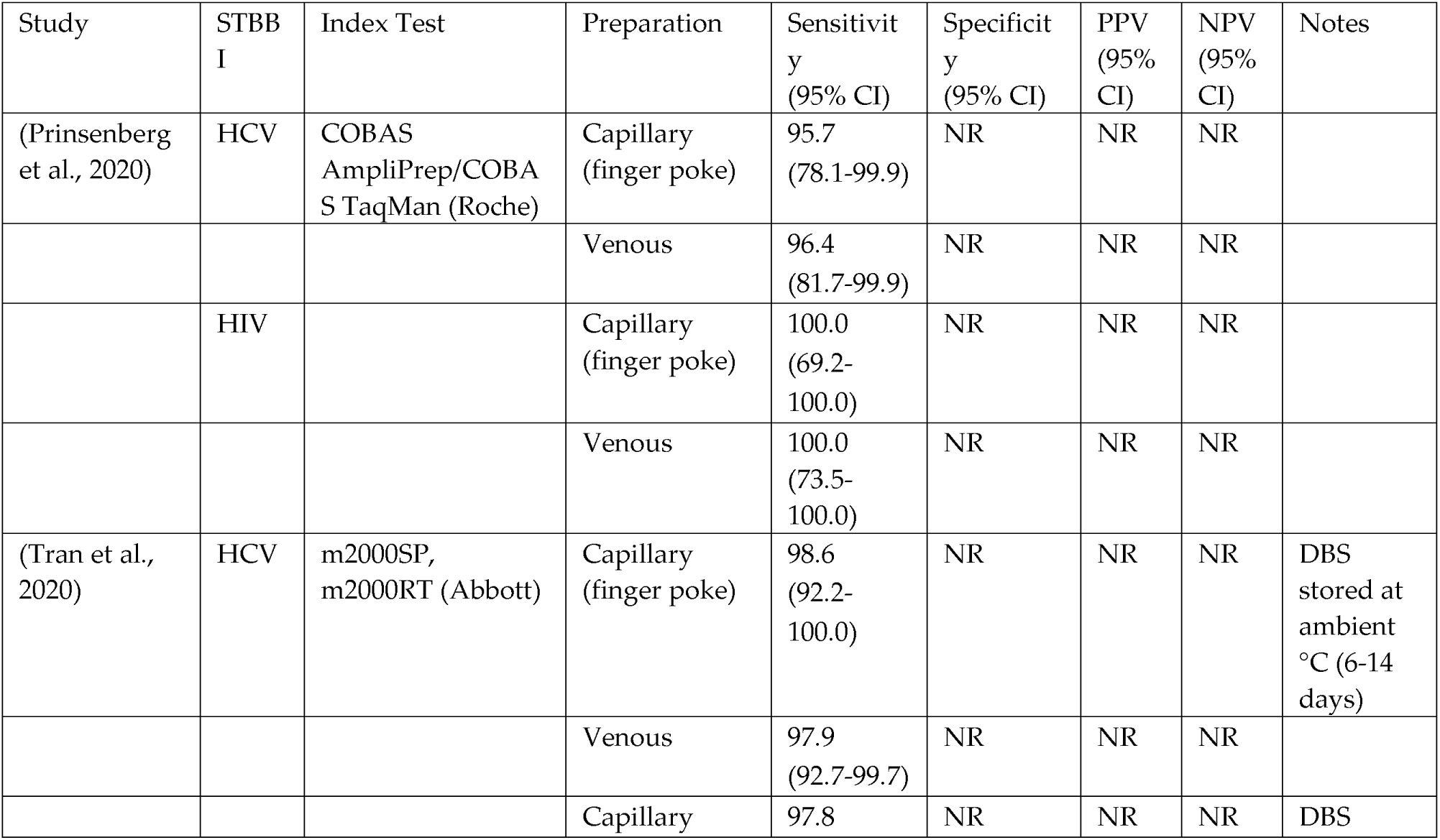
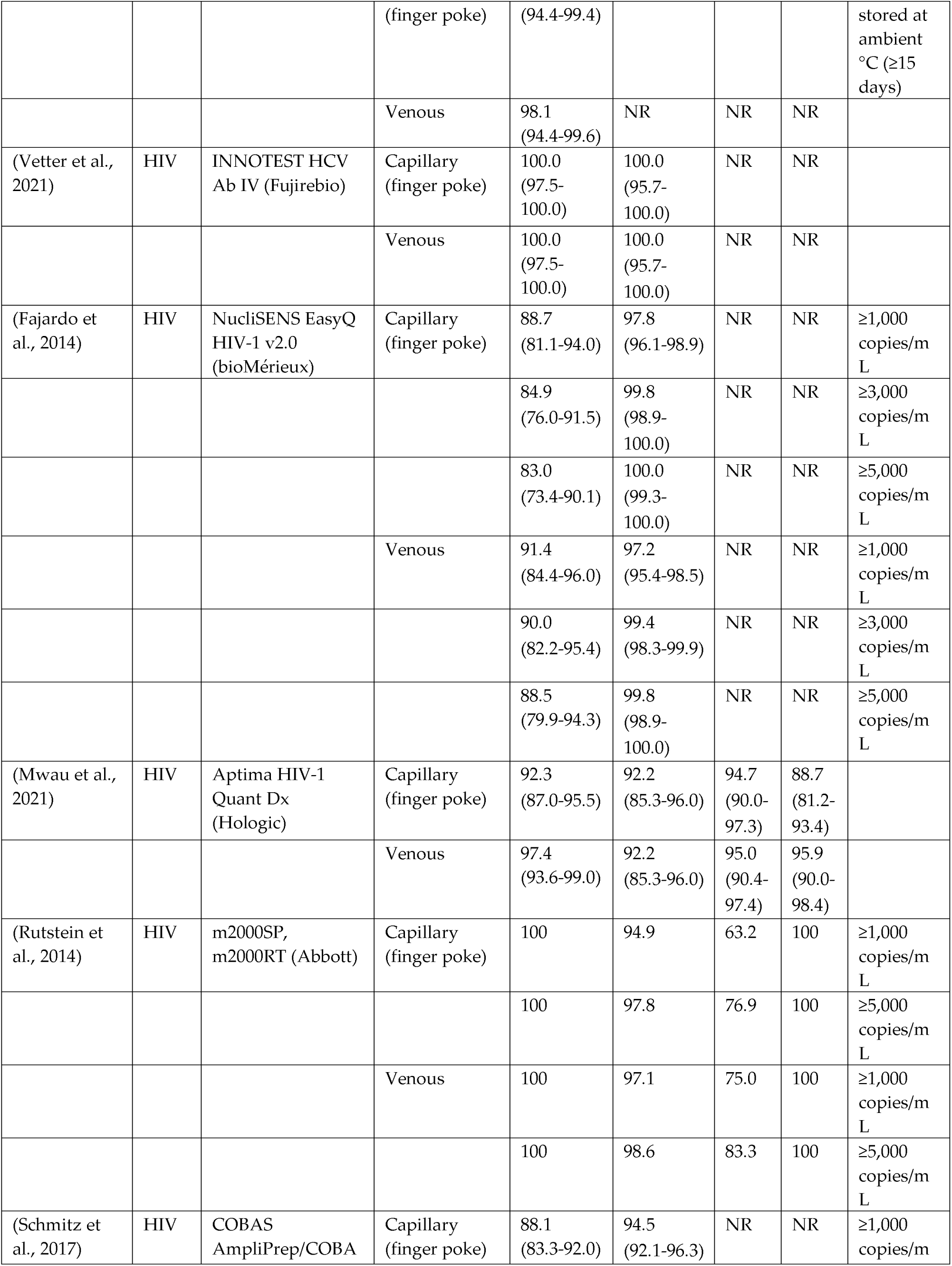
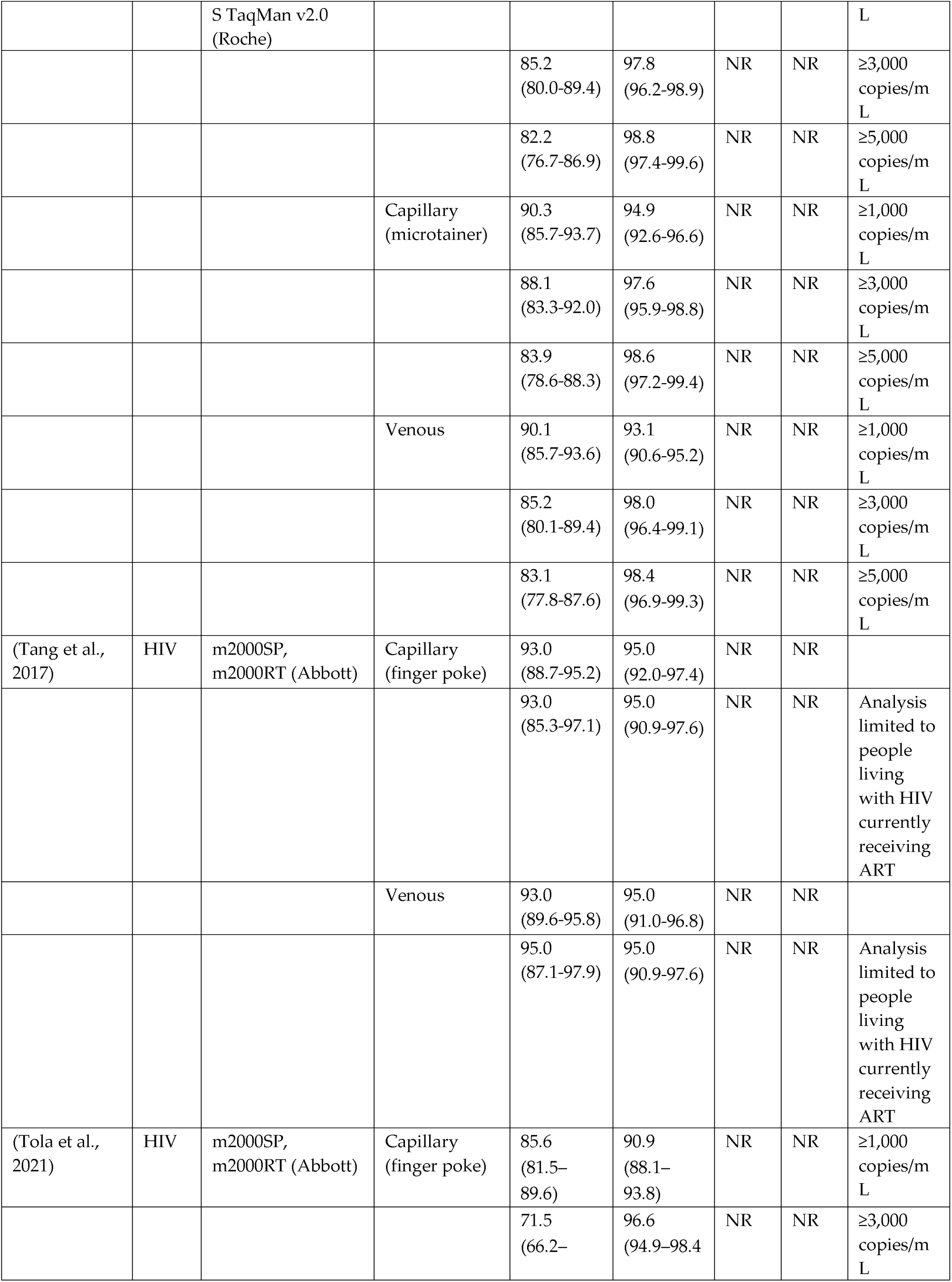
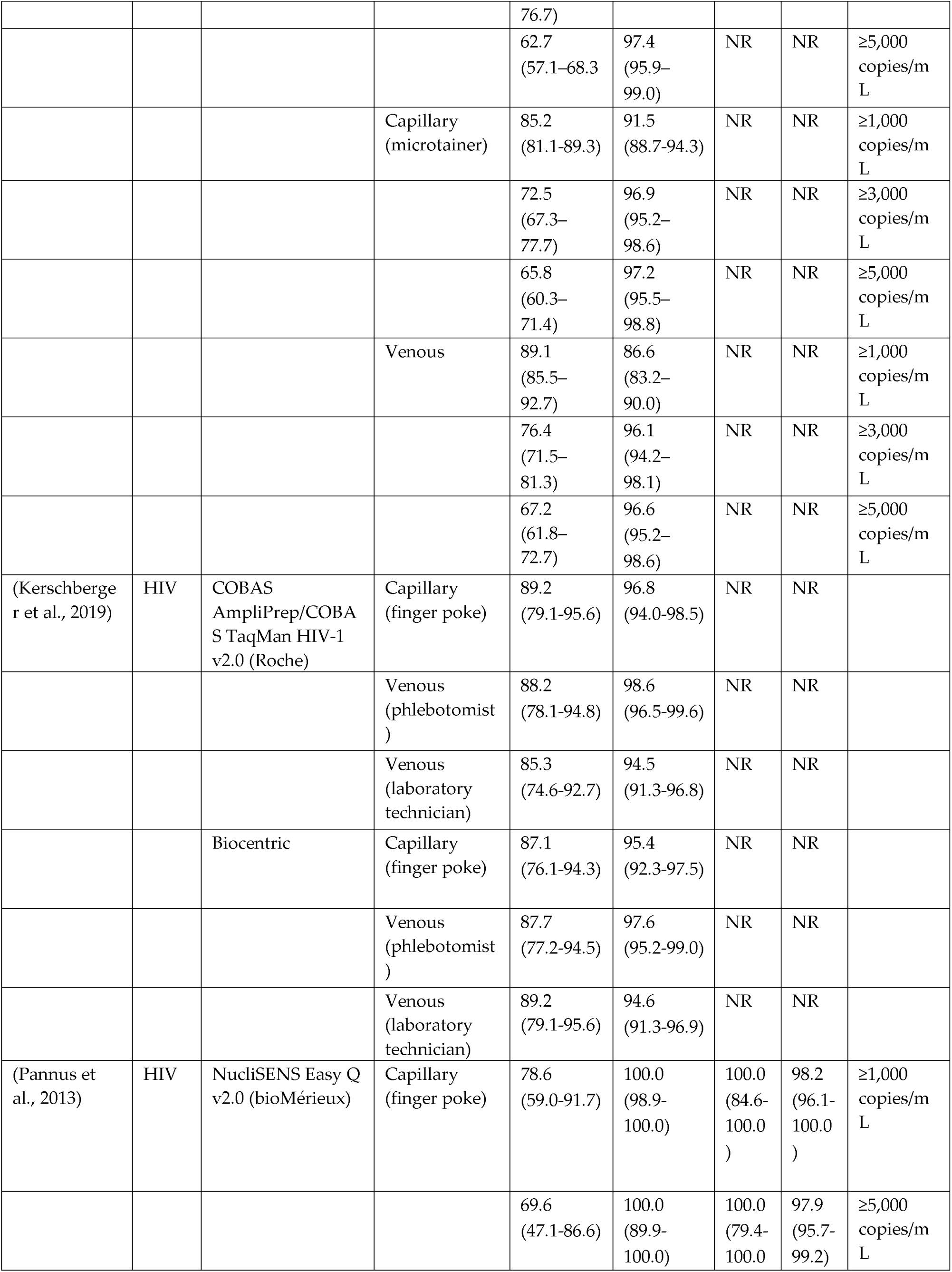
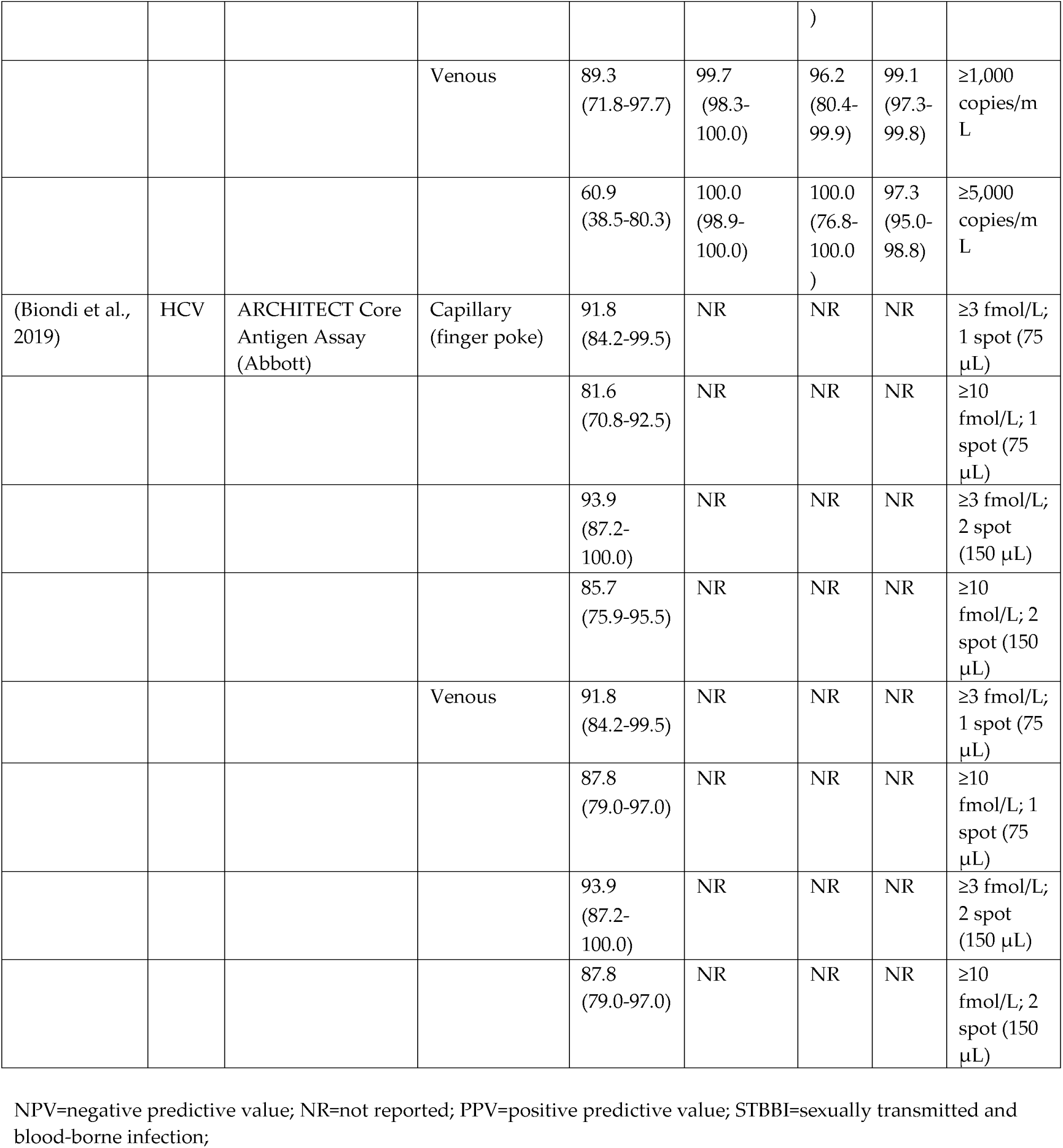
Studies comparing DBS prepared from venous and capillary blood

### Dual Infections

The presence of dual infections may influence test performance depending on the STBBI of interest (Table 4). This was particularly evident when testing for HBV and HCV on DBS collected from individuals living with HIV. Flores et al. (Flores, Cruz, Potsch, et al., 2017) reported lower sensitivity and specificity in persons living with HIV compared to those without HIV for the detection of both HBV surface antigens (HBsAg) and anti-hepatitis B core total antibodies (Anti-HBc) on the Elecsys platform (Roche; Table 4). Similar findings have also been observed in HCV testing. De Crignis et al. (De Crignis, Re, Cimatti, Zecchi, & Gibellini, 2010), Saludes et al. (Saludes et al., 2018), and Flores et al. (Flores et al., 2018) all reported reduced HCV test sensitivity in people living with HIV (Table 3). In contrast, Flores et al. (Flores et al., 2021) reported increased HCV test performance in people living with HIV compared to those who were not, but chronic kidney disease among study participants may have confounded these findings. Nonetheless, the presence of potential co-infections should be taken into consideration when validating a test for use with DBS specimens.

**Table 4.**
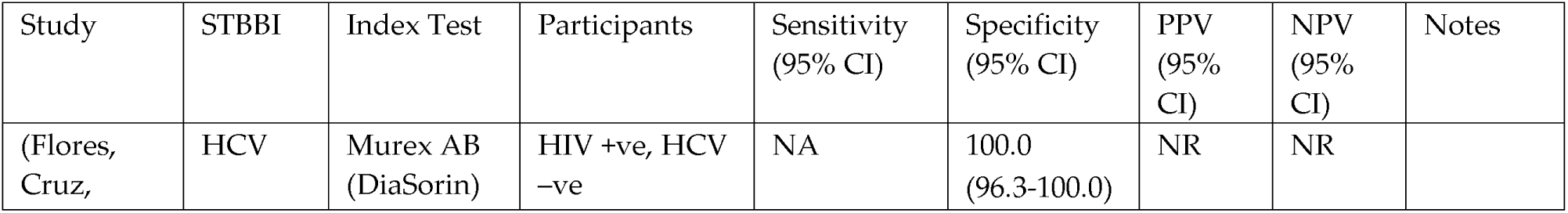
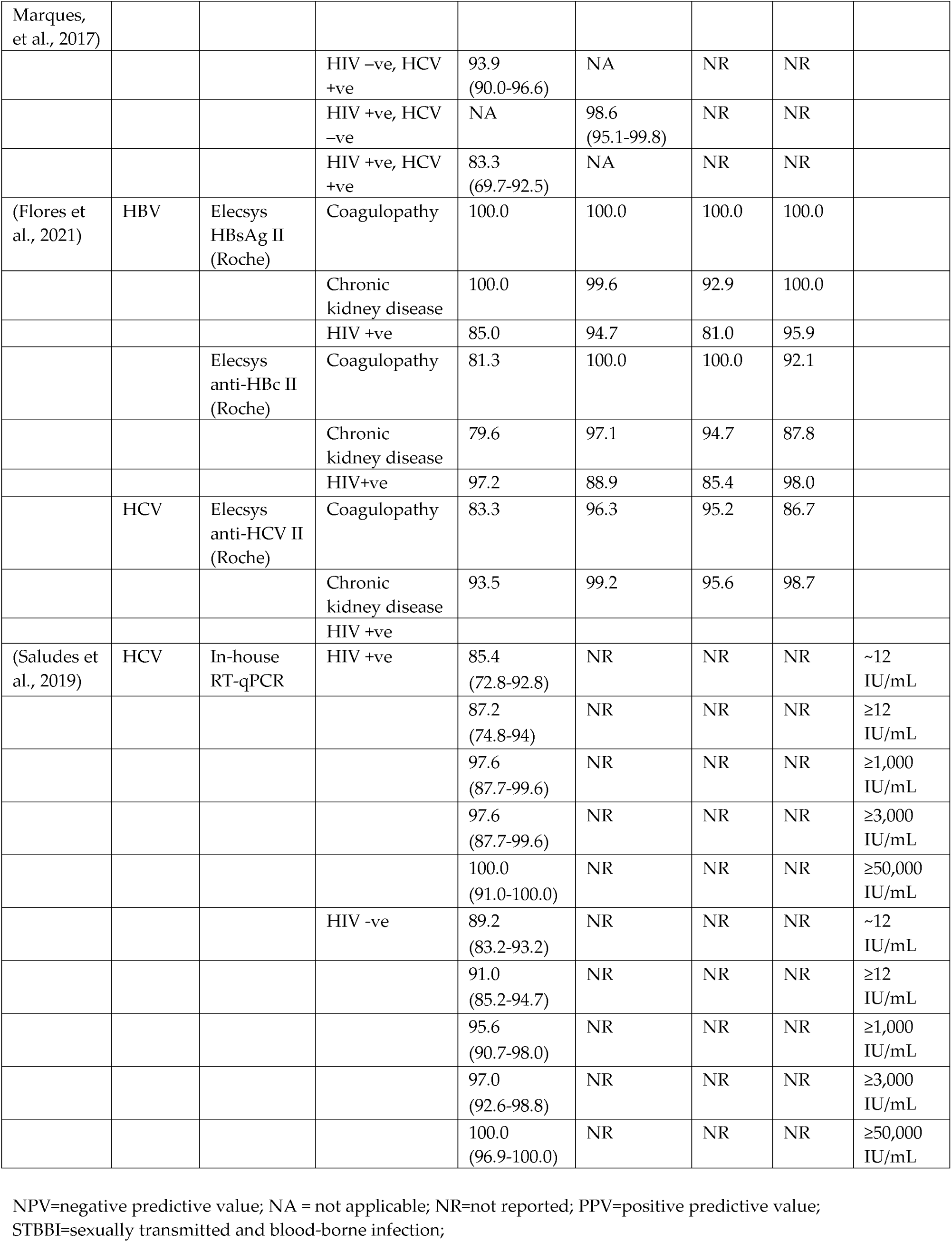
Studies assessing test performance on DBS specimens collected from participants with co-infections

### Antiretroviral Therapy

In studies which included people living with HIV, assay performance was typically assessed in patients undergoing antiretroviral therapy (ART) in comparison to ART-naïve patients to detect treatment failure (Table 5). Balinda et al. (Balinda et al., 2016) reported differences in the performance of an in-house HIV RT-qPCR assay in participants living with HIV who were undergoing ART compared to those who were ART-naïve (79.4% sensitivity, 54.5% specificity), with the highest performance observed in patients who had undergone ART for longer periods (12 – 36 months; 88.9% sensitivity, 98.1% specificity). Similarly, Taeib et al. (Taieb et al., 2018) reported greater specificity in patients on ART for ≥6 months compared to those on ART for <6 months but, found no difference in sensitivity.

**Table 5.** Studies assessing test performance on DBS specimens collected from participants undergoing ART and ART-naïve participants

| Study | STBBI | Index Test | Participants | Sensitivity<br>(95% CI) | Specificity<br>(95% CI) | PPV<br>(95%<br>CI) | NPV<br>(95%<br>CI) | Notes |
| --- | --- | --- | --- | --- | --- | --- | --- | --- |
| (P. Alvarez et al., 2015) | HIV | COBAS AmpliPrep/COBAS TaqMan HIV-1 Quantitative Test v2.0 (Roche) | ART-naïve | 94.7<br>(74.0-99.9) | 90.5<br>(77.4-97.3) | NR | NR |  |
|  |  |  | Undergoing ART | 96.2<br>(86.8-99.5) | 90.5<br>(80.4-96.4) | NR | NR |  |
|  |  |  | All | 95.8<br>(88.1-99.1) | 89.5<br>(82.0-94.7) | NR | NR |  |
|  |  | Versant HIV-1 RNA 1.0 (Siemens) | ART-naïve | 84.2<br>(60.4-96.6) | 100.0<br>(91.0-100.0) | NR | NR |  |
|  |  |  | Undergoing ART | 64.7<br>(50.4-77.6) | 100.0<br>(93.4-100.0) | NR | NR |  |
|  |  |  | All | 70.0<br>(57.9-80.4) | 100.0<br>(96.1-100.0) | NR | NR |  |
| (Balinda et al., 2016) | HIV | In-house RT-qPCR | ART-naïve | 79.4 | 54.5 | 89.0 | 36.4 |  |
|  |  |  | Undergoing ART (>6 months) | 75.7 | 95.5 | 65.1 | 97.3 |  |
|  |  |  | Undergoing ART (12-36 months) | 88.9 | 98.1 | 72.7 | 99.3 |  |
| (Ondoa et al., 2014) |  | COBAS AmpliPrep/COBAS TaqMan HIV-1 Quantitative Test v2.0 (Roche) | ART-naïve | 61.9 | 99.0 | NR | NR | ≥5,000 copies/mL |
|  |  |  | Undergoing ART | 9.0 | 100.0 | NR | NR | ≥5,000 copies/mL |
| (Taieb et al., 2018) |  | m2000SP, m2000RT (Abbott) | Undergoing ART (<6 months) | 87.5<br>(47.3-99.7) | 87.1<br>(70.1-96.4) | NR | NR |  |
|  |  |  | Undergoing ART (≥6 months) | 85.2<br>(66.3-95.6) | 99.0<br>(94.3-100.0) | NR | NR |  |
|  |  |  | All | 90.1<br>(80.7-95.9) | 96.2<br>(91.4-98.8) | NR | NR |  |

### Quality Assessment

All 168 included studies were assessed for quality using the QUADAS-2 tool (Fig. 3). The overall quality of studies was low, with high scores in risk of bias and applicability. Risk of bias was found to be high across most domains, with 117 studies (69.6%) rated as having high risk of bias in patient selection, 85 (50.6%) as high risk of bias for the index test, and 134 (79.8%) as high risk of bias for the reference standard. In addition, a small number (*n*=12; 7.1%) of studies were rated as high risk of bias related to flow and timing. Similarly, a significant proportion of studies were rated as high concern regarding the applicability of the study to the research question. The proportion of studies rated as high concern regarding the applicability of population, index test, and reference standard was found to be 23.2%, 47.6%, and 47.6%, respectively.

**Figure 3.**
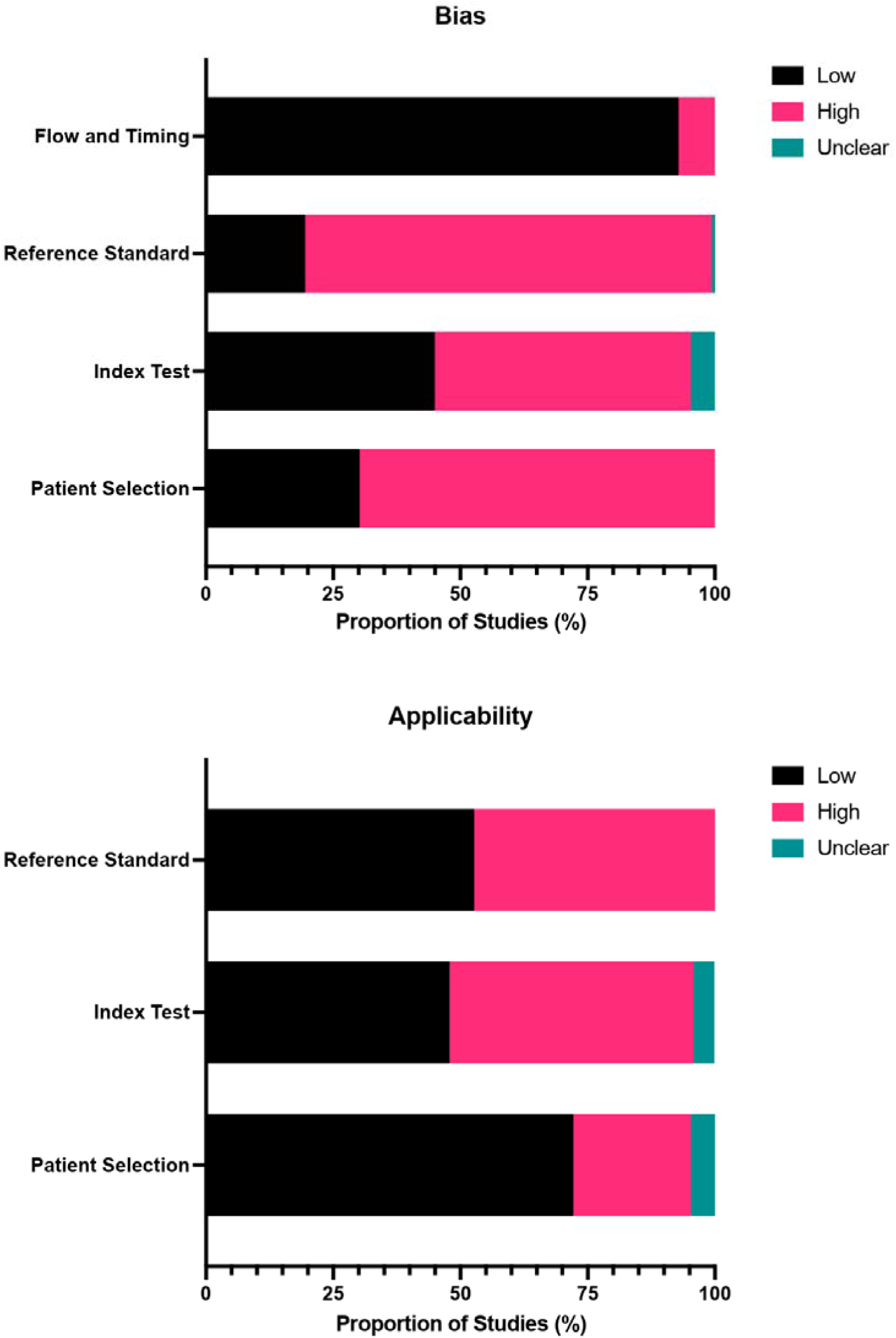
Proportion of studies with low, high, or unclear risk of bias and concerns regarding applicability based on the QUADAS-2 tool

## Discussion

This review describes the validity of using DBS specimens for STBBI testing. DBS specimens showed high sensitivity compared to the reference standard of plasma or serum in the included studies encompassing a wide range of experimental conditions, demonstrating a promising opportunity for the adoption of DBS specimens in STBBI testing globally. The high sensitivity and specificity observed across studies provide evidence for the suitability of DBS for the surveillance of STBBIs, including those not routinely detected in DBS specimens, such as HAV, HTLV and syphilis.

DBS specimens offer a promising alternative to plasma and serum samples for STBBI testing, though assay performance depends on several factors. We identified multiple factors which could influence test performance with DBS specimens. Adjustment of test cut-off values for DBS specimens improved overall performance (García-Cisneros et al., 2019), making it imperative for individual laboratories (or manufacturers) to validate specific cut-off values for use with DBS specimens. The presence of dual infections within a population also influenced assay performance (Flores, Cruz, Potsch, et al., 2017), resulting in decreased sensitivity, especially for HBV and HCV testing in participants living with HIV. This finding is relevant for future test development and validation for future surveillance in key populations where dual infections may be more prevalent and in geographies which experience overlapping STBBI burdens (Shayan, Nazari, & Kiwanuka, 2021). Approaches to DBS preparation (venous versus capillary blood) also affected test performance (Prinsenberg et al., 2020). Although DBS prepared from venous blood appears to offer better test performance (especially for HIV nucleic acid testing), DBS prepared from capillary blood still offer excellent test performance and reflects more closely “real-world” DBS collection, especially considering self-collection. Enhanced reporting of DBS validation studies could lead to further improvements in the performance of DBS testing using capillary blood. Finally, test performance may fluctuate depending on the duration of ART among people living with HIV. Our review supports the validity of using DBS specimens for detecting certain STBBIs. DBS specimens perform well particularly well in HIV and HCV testing, though no conclusions can be made for chlamydia or gonorrhoea surveillance as no studies investigated DBS testing with either pathogen. However, test performance relies on several experimental conditions, and standardized approaches to reporting DBS experiments should be adopted moving forward to ensure internal and external test validity.

Due to considerable heterogeneity observed across studies, we could not conduct a meta-analysis. General poor reporting of experimental conditions also created challenges in directly comparing studies – this was also reflected in our quality assessment using the QUADAS-2 tool. There is currently no consensus on how DBS studies should be reported. We propose that new standardized guidelines for reporting DBS experiments should be developed and implemented similarly to the MIQE guidelines for quantitative real-time PCR experiments (Bustin et al., 2009) or the STARD checklist (Bossuyt et al., 2015). Reporting guidelines should include the minimum information required to evaluate DBS studies, including information on DBS preparation (venous or capillary blood), drying and storage conditions, type of filter paper used, and DBS elution protocols (for example volume, type of buffer, and agitation conditions). This would allow for a more direct comparison of studies and assist in conducting robust meta-analyses to further investigate the validity of DBS specimens for STBBI testing. Though we did not record storage temperature and humidity in this review, these factors have been examined by (Amini et al., 2021) in a systematic review on on reliability of antibody measurement in DBS specimens. Though their objective differs from our review and includes fewer papers (n=40), the similar observations of heterogeneity in experimental conditions and reporting further our observation that it is necessary to record a large number of influential variables to allow for standardized assessment of studies.

While treatment failure was not explicitly part of our objective, it is an important component of surveillance. More specifically, it can provide insights into the cascade of care and progress toward HIV and HCV elimination targets (Dore & Bajis, 2021; UNAIDS, 2014). Viral load testing alone is not sufficient for the purposes of surveillance as persons undergoing treatment may present undetectable viral loads, and therefore must be used in combination with serological assays to establish disease status. We decided to include studies investigating viral load with DBS specimens as viral load data still provide valuable insight into the cascade of care or other metrics towards elimination. It should be noted that the LOD for most HIV tests included in this review was determined to be approximately 800 copies/mL. The WHO defines virological suppression as <1,000 copies/mL, which should be reliably detected by most assays in this review (Patricia Alvarez et al., 2015; Erba et al., 2015). This finding is in agreement with a recent systematic review conducted by Vojnov et al. (2022) which concluded DBS specimens are suitable for HIV viral load testing at the treatment failure threshold of 1,000 copies/mL (Vojnov et al., 2022)[X]. Although we consider DBS specimens suitable for certain STBBI surveillance, careful attention should be paid to the LOD of each assay under consideration and consider definitions of treatment failure for the STBBI of interest.

Limitations of this review include the unavailability of any studies on DBS testing for chlamydia and gonorrhoea. We limited our biological specimen of interest to blood, and therefore did not include any papers on dried urine spots for chlamydia or gonorrhoea detection. Additionally, we were unable to conduct a meta-analysis as a result of the large amount of heterogeneity between studies. Finally, we did not examine acceptability of DBS testing among participants versus traditional methods, as this is beyond the scope of this review and warrants its own investigation.

## Conclusion

Over the course of the COVID-19 pandemic, the use of DBS specimens increased in popularity as many studies adopted them for serological studies (Cholette et al., 2022; Madhi et al., 2022; Miesse et al., 2022; Wong et al., 2022). We anticipate that this increase in DBS usage will carry forward and result in a greater number of studies using DBS specimens for STBBI testing. This review highlights the validity of DBS sampling for use in STBBI testing and identifies several key factors which influence assay performance. The use of DBS sampling presents a promising opportunity for use in STBBI surveillance in remote and isolated communities in Canada. In addition, it can benefit STBBI screening in other resource constrained settings.

## Data Availability

All data generated or analysed during this study are included in this published article (and its Supplementary Information files).

## Acknowledgments

We thank Janice Linton (Indigenous Health Librarian & Liaison Librarian for Community Health Sciences at University of Manitoba, Winnipeg, Canada) for generously giving her time and assistance with the initial literature search.

## Notes

**Declarations Funding:** KMS is supported by a Tier 2 Canada Research Chair in Integrated Knowledge Translation in Rehabilitation Sciences.

**Conflict of interest:** The authors declare no competing interests.

### Competing Interest Statement

The authors have declared no competing interest.

### Funding Statement

This study did not receive any funding.
Kathryn M. Sibley is supported by a Tier 2 Canada Research Chair in Integrated Knowledge Translation in Rehabilitation Sciences.

