## Supplementary File 1 for "Validity of Dried Blood Spot Testing for Sexually Transmitted and Blood-Borne Infections: A Narrative Systematic Review"

*Search strategy MeSH terms and keywords*

(“Hepatitis A” or “HAV” or “Hepatitis B” or “HBV” or “Hepatitis C” or “HCV” or “Non-a, non-b hepatitis” or “Human immunodeficiency virus” or “HIV” or “Lymphadenopathy-associated virus” or “LAV” or “Herpes simplex” or “Herpesvirus” or “HSV” or “Human T-lymphotropic virus” or “HTLV” or “Human papilloma virus” or “Human papillomavirus” or “HPV” or “Chlamydia” or “Chlamydia trachomatis” or “Gonorrh*” or “Neisseria gonorrhoeae” or “Syphilis” or “Treponema pallidum” or “Hepatitis, Viral, Human” or “Parenterally-transmitted non-A, non-B hepatitis” or “HIV-1” or “HIV-2” or “Herpes virus 1, Human” or “Herpes virus 2, Human”, or “Simplexvirus” or “Human T-lymphotropic virus 1” or “Human T-lymphotropic virus 2” or “Human T-lymphotropic virus 3” or “Deltaretrovirus” or “Alphapapillomavirus” or “Sexually transmitted diseases, bacterial” or “Sexually transmitted diseases, viral” or “Chlamydia infections”) and (“Dried blood spot” or “Dried blood” or “Dried plasma” or “Dried plasma spot” or “Filter paper” or “DBS” or “Ahlstrom” or “Munkell” or “Whatman” or “Guthrie” or “Hemaspot*” or “Dried blood spot testing”) and (“Sensitivity” or “Specificity” or “Accuracy” or “Positive predictive value” or “PPV” or “Negative predictive value” or “NPV” or “Limit of detection” or “LOD” or “Limit of quantification” or “LOQ” or “Likelihood raio” or “Sensitivity and Specificity” or “Predictive value of tests” or “ROC curve” or “Data accuracy” or “Reproducibility of results”)
